# Survival rate of positive peritoneal cytology in endometrial cancer; a systematic review and meta-analysis

**DOI:** 10.1101/2022.04.21.22274129

**Authors:** Hamidreza Dehghan, Fariba Binesh, Mohammad Taghi Moravej, Ali Zare Dehnavi, Hojat Dehghanbanadaki, Safiyehsadat Heydari, Maryam Nikfard, Mehrdad Mansouri

## Abstract

**Background:** The impact of positive peritoneal cytology on survival rate of endometrial cancer patients in different stages and histopathology is still controversial. We performed a systematic review and meta-analysis to investigate the influence of positive peritoneal cytology (PPC) on survival rate of patients with endometrial carcinoma.

**Methods:** A systematic literature search of PubMed, Embase, Scopus, and Cochrane databases was conducted up to November 24, 2020. The quality of included studies was evaluated by Quality in prognosis study (QUIPS) tool.

**Results:** Initially, 3014 articles were found, of which 65 met the inclusion criteria for qualitative analysis and 27 studies on 75897 patients with endometrial cancer were included in the meta-analysis. PPC was associated with a lower overall survival in endometrial cancer (HR= 2.102; 95% CI:1.629-2.711; P< 0.001). The findings also identified PPC as an independent prognostic factor for both disease-free survival (HR= 3.052; 95% CI: 2.348-2967; P< 0.001) and cancer specific survival (HR= 3.461; 95% CI: 2.280-5.254; P< 0.001). In addition, we meta-analyzed the studies in 21 subgroups based on staging and histopathology of the endometrial cancer which all identified PPC as a non-prognostic factor for cancer of endometrium.

**Conclusion:** PPC is an independent prognostic factor for endometrial cancer survival rate in all staging and histopathologic subgroups.

## Introduction

Endometrial cancer is considered the most common gynecological cancer in industrialized countries and the second most common in developing countries (1). Its incidence has increased steadily since 1990, with the highest growth rate for countries with a high sociodemographic index (SDI) (2). Despite the fact that approximately 75% of endometrial cancer cases are diagnosed in early stages, the annual estimated rate of endometrial cancer mortality is increasing rapidly at a rate of 1-2%(3,4). Most of the mortality occurs in advance disease, which highlights the importance of prognostic factors in endometrial carcinoma and early detection of cases (5).

The importance of numerous prognostic factors has been thoroughly investigated, and multiple prognostic factors have been identified, including age, parity, tumor grade, and cancer stage (6,7). Of these factors, tumor stage is the most powerful prognostic parameter and can provide a useful tool for predicting patients’ outcomes and facilitating recommendation of proper treatment options (3,8,9). Endometrial cancer staging was previously done clinically until 1988, when the International Federation of Gynecology and Obstetrics (FIGO) introduced a surgical staging system(10). In 2009, the FIGO staging system for endometrial cancer was revised, and peritoneal cytology was removed as a staging criteria (7,11). Despite the omission of peritoneal cytology from the staging system, several studies have shown contradictory results, and positive peritoneal cytology has been found useful for evaluation of prognosis and prediction of survival rate (12– 14).

Despite all previous investigations, the role of positive peritoneal cytology in endometrial cancer staging is still a matter of debate, and excluding positive peritoneal cytology from the staging system may mislead prognosis estimations in these patients, resulting in under treatment and an increasing mortality rate (11). In this study, we have systematically reviewed and meta-analyzed the existing literature on the effect of positive peritoneal cytology on the survival rate of patients with endometrial cancer to clarify the clinical importance of PPC in endometrial carcinoma.

## Materials and methods

### Protocol registration

This systematic review and meta-analysis was conducted using the Preferred Reporting Items for Systematic Reviews and Meta-Analyses (PRISMA) (15). Furthermore, the study protocol has been registered in PROSPERO with the code of CRD42018103587 and published in detail previously (16).

### Search strategy and screening

Initially, on October 30, 2018, we searched PubMed, Embase, Scopus, and Cochrane databases for all original studies without time and language limitations. Following the initial search, an alarm was set in each database to notify us about new manuscripts, and new studies were reviewed before the final analysis (November 24, 2020). The following terms were used for searching the databases: endometrial cancer, endometrial carcinoma, endometrial neoplasm, endometrial tumor, peritoneal washing, peritoneal lavages, peritoneal irrigations, peritoneal cytology, peritoneal lavage cytology, positive peritoneal washing, and positive peritoneal cytology. Two investigators independently assessed the title, abstract, and full-text of the retrieved papers, and any disagreements were resolved with a third investigator. Then, the included articles were surveyed for data extraction. In the cases where the full text or required data for conducting the systematic review and meta-analysis was missing, up to three emails were sent to the corresponding author of these papers at two-week intervals, and they were asked for the missing data.

### Inclusion and exclusion criteria

The inclusion criteria for study selection were: all original studies on endometrial cancer patients who underwent peritoneal washing cytology and were also assessed for the survival. Papers which the full text of the manuscript was not provided were excluded. Studies with incomplete or missing medical data which failed to meet the requirements for our checklist were also excluded.

### Data extraction and measured outcomes

Two authors independently extracted data from the included papers using the pre-designed data extraction form, and disagreements were discussed and resolved through consultation with a third investigator. The data extraction check list consisted of the following items: bibliographic data (article title, paper’s first author name, publication year, journal name, year of publication, country, and type of study), study population characteristics (number of patients, age, tumor grade and stage, histopathology findings, and type of treatment), number of positive peritoneal cytology, hazard ratio (HR) and its dispersion for overall survival (OS), disease -free survival (DFS), and cancer specific survival (CSS).

The main measured outcomes were overall survival (OS), defined as the time which begins at diagnosis and up to the time of death due to any cause, disease free survival (DFS), defined as the time from diagnosis to recurrence of the disease, and cancer specific survival (CSS), defined as the time from diagnosis of the disease to death from that disease.

### Quality assessment

The Quality in prognosis study (QUIPS) tool (17,18) was used to evaluate the quality of included studies and to measure the bias of the papers.

### Statistical analysis

STATA 16 was used for statistical analyses. The logarithm of HR and standard error of logarithm of HR were calculated, and forest plots were generated using fixed-effects model when there was no substantial heterogeneity (I2< 50%); otherwise random-effect model was used. Heterogeneity was assessed through Cochran’s Q test and I square index. The funnel plot and Egger test was used to measure publication bias. Subgroup analysis was performed based on the staging method (FIGO 1988, FIGO 2009), the studied stage (low stage, high stage, and all stages together), and the investigated histology (Type 1 histology, Type 2 histology, and all histologies together).

## Results

### Study selection

A total of 3014 articles were identified during the initial search in four databases. After removing the duplicates, a total of 2142 articles were obtained, of which 138 reached the full-text assessment stage. Of these, 59 articles were excluded regarding eligibility criteria, and 21 articles were not included due to unavailability of full-texts (Figure 1). Finally, 65 papers were included for qualitative analysis which were retrospective observational studies and were published from 1981 to 2020 (7,9,11,14,19–77). The details of these studies are also presented in Appendix 1.

**Figure1.**
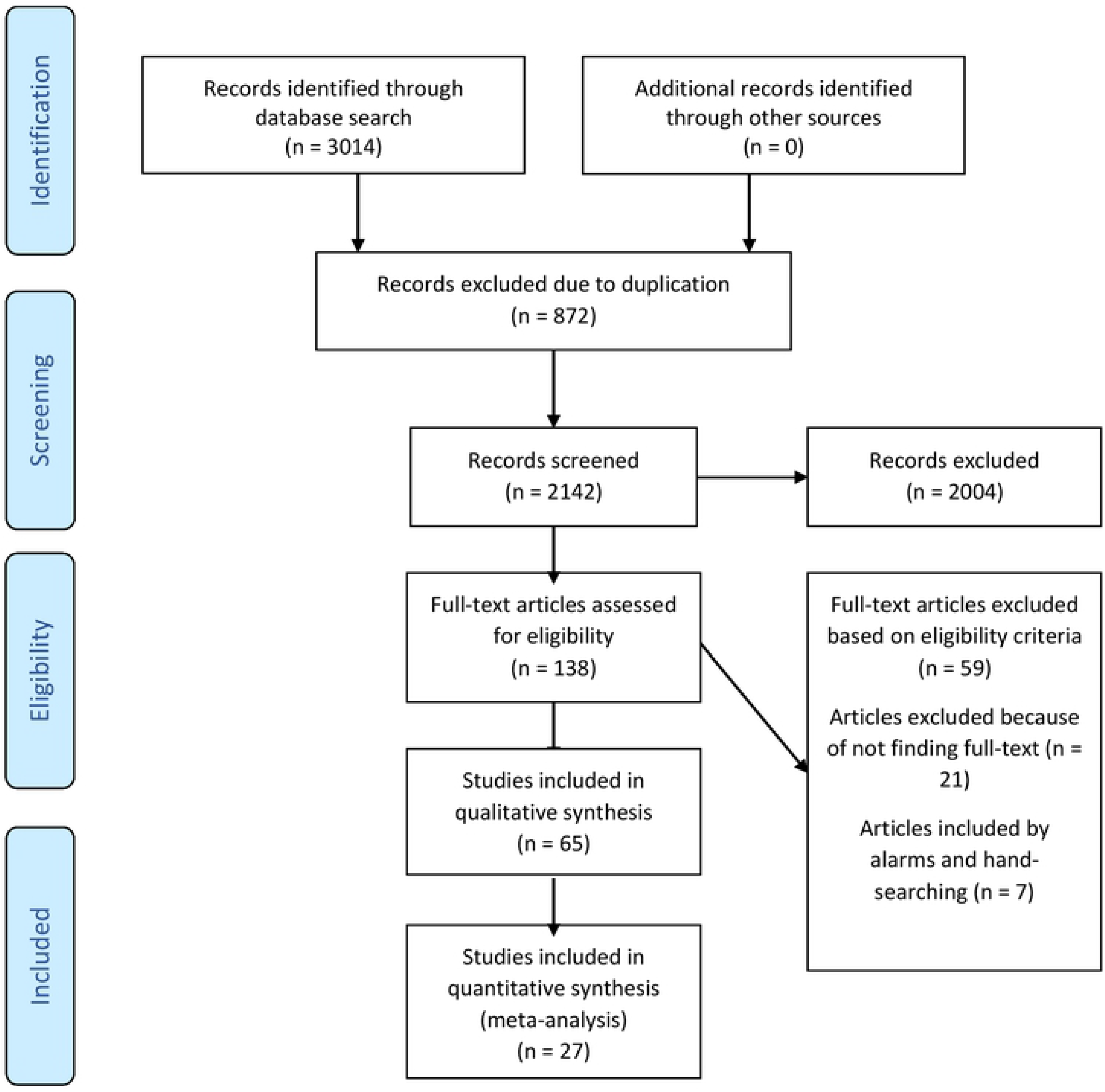
PRISMA flow diagram. Start date: Oct 30, 2018; End date: Nov 24, 2020.

### Characteristics of included studies

Of these 65 studies, 27 papers met the criteria for meta-analysis, which included an overall number of 75897 patients. Of the 27 papers, 16 studies (11,13,21,22,24,35,36,43,49,54,58,62,68,70,73,77) included OS data, 18 had DFS data(14,21,62,64,68,70,72,74,77,78,25,36,39,43,44,49,53,58), and 7 had CSS(7)(19)(33)(44)(45)(64)(73). The quality assessment (QUIPS) of included studies is summarized in table 1.

**Table1.**
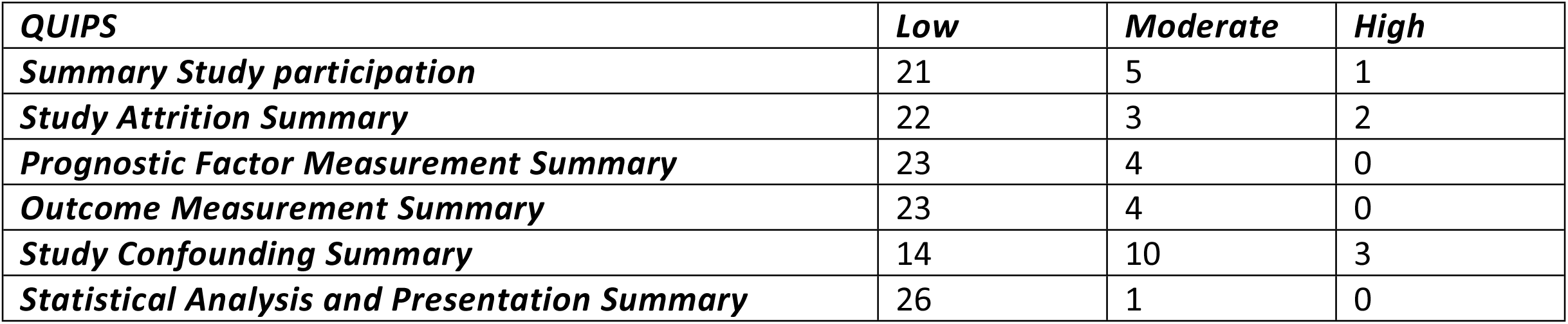
Summary of included studies’ QUIPS assessment

### Quantitative analysis

Results of the meta-analysis on the overall survival rate for 16 papers are shown in Figure 2. Figures 3 and 4 present the results of meta-analysis on DFS and CSS, respectively. The findings of these analyses can be summarized as follows:

OS: HR: 2.102(CI (95%):1.629-2.711, P: 0.000, Z: 6.913)

DFS: HR: 3.052(CI (95%):2.348-2967, P: 0.000, Z: 8.342)

CSS: HR: 3.461(CI (95%):2.280-5.254, P: 0.000, Z: 5.831)

**Figure2.**
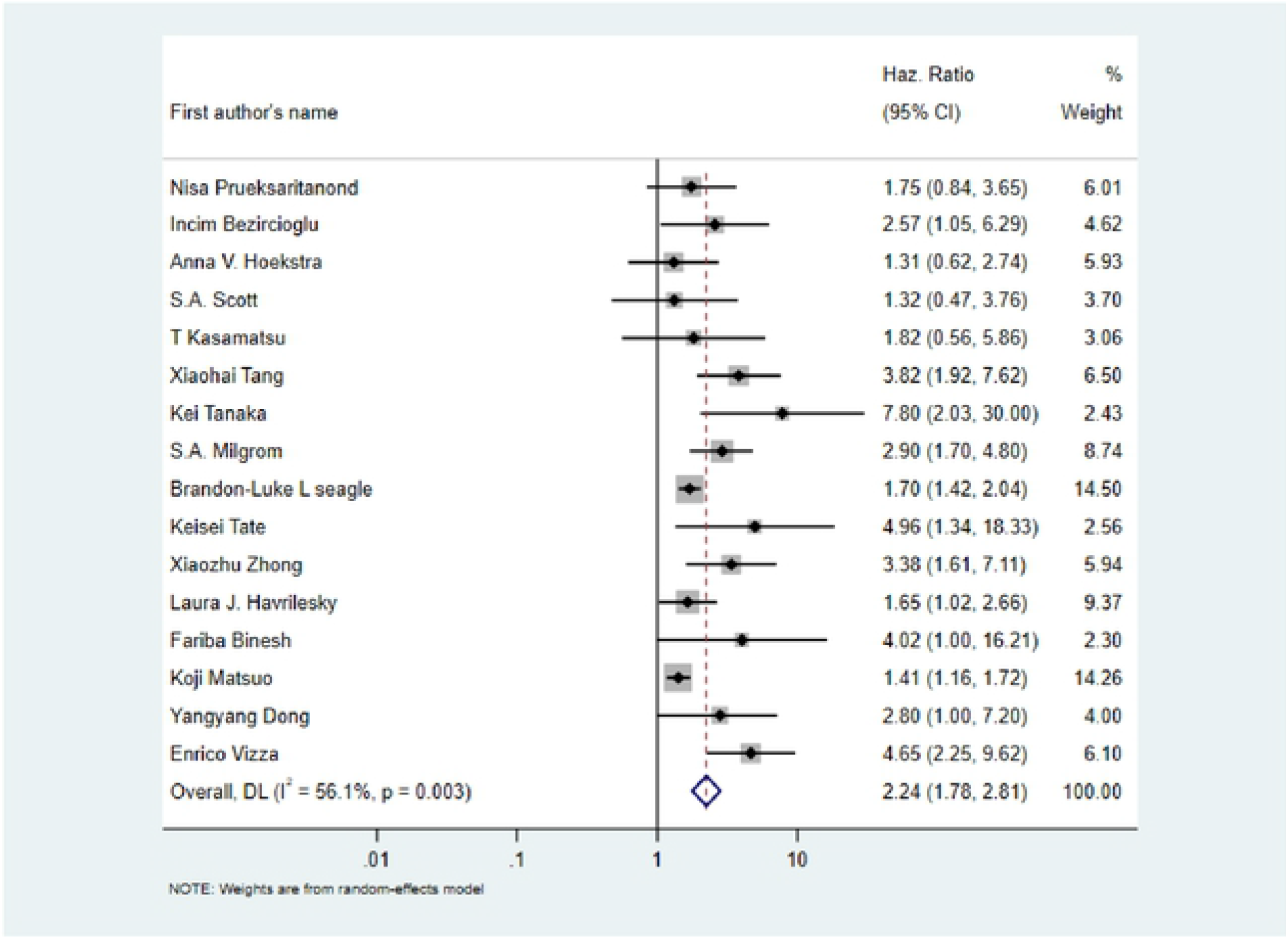
Forest Plot of the articles analyzing OS.

**Figure3.**
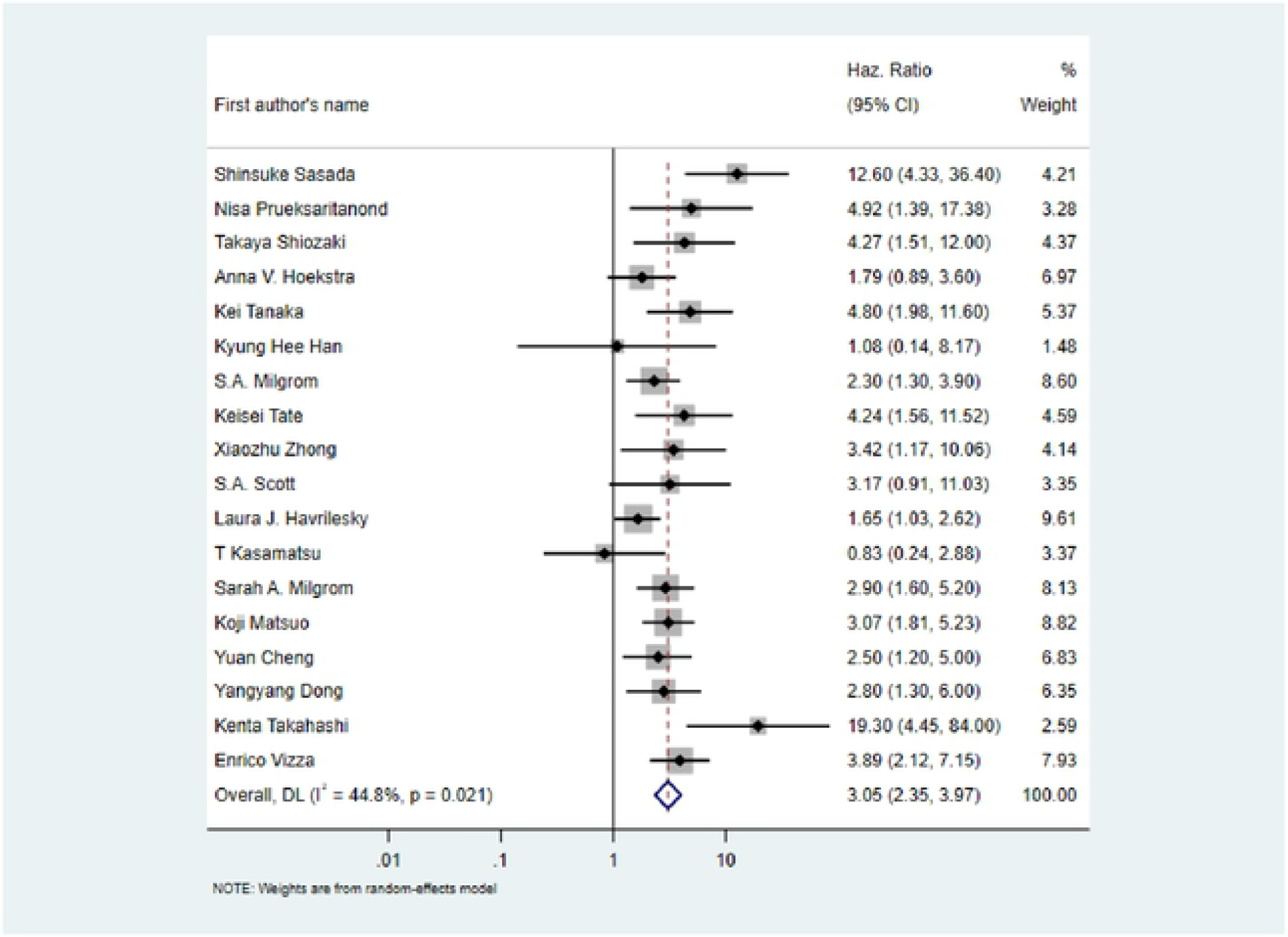
Forest Plot of the articles analyzing DFS.

**Figure4.**
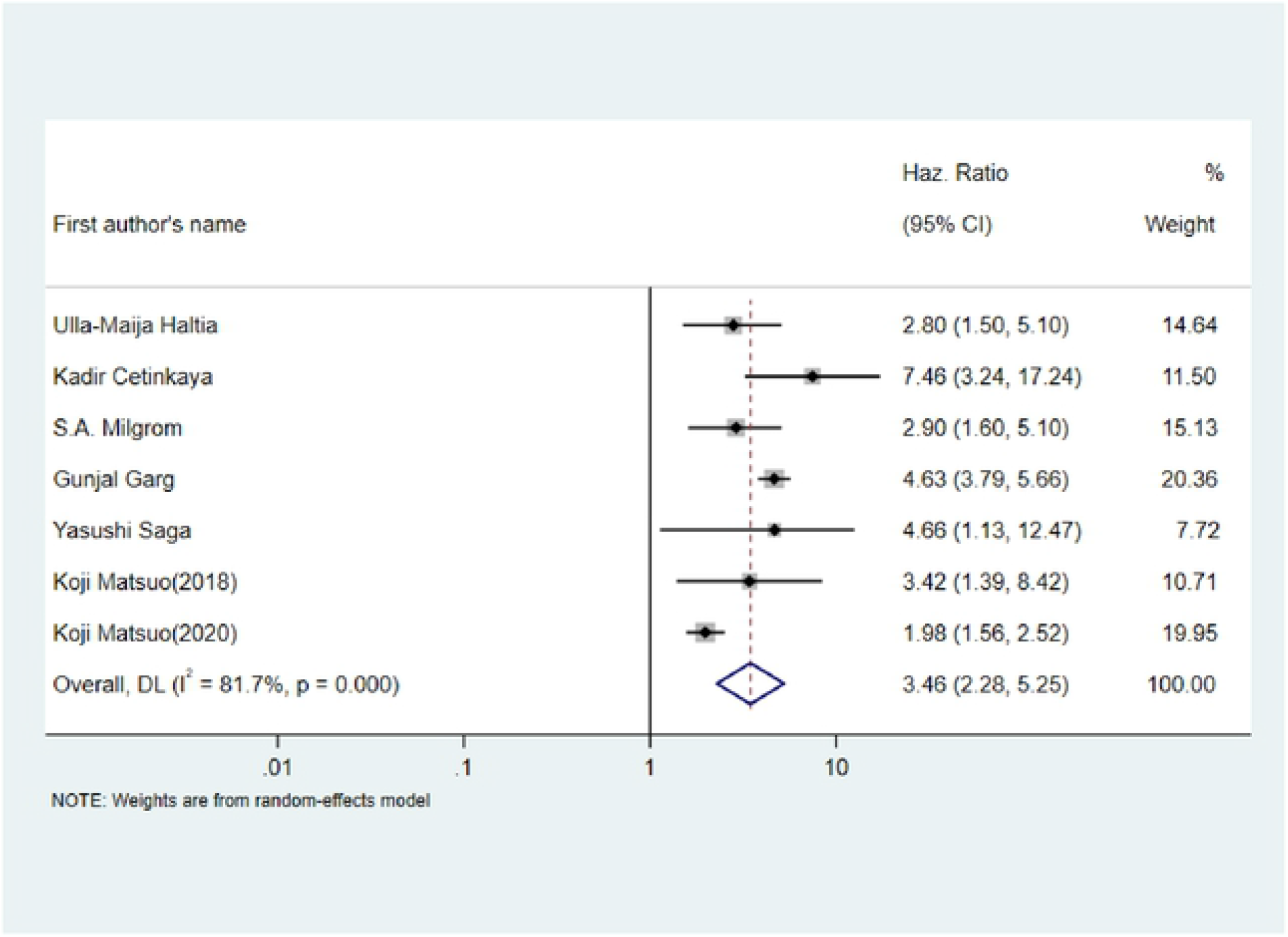
Forest Plot of the articles analyzing CSS.

### Subgroup Meta-analysis

We also analyzed the outcome of endometrial cancer patients in different subgroups based on staging system and histopathology. The details of subgroups’ analysis findings are reported in table 2. The results of subgroup analysis indicated that of the total 21 subgroups, 12 had low heterogeneity (<50%), 7 had moderate heterogeneity (75%–50%), and 2 showed high heterogeneity (>75%). Detailed results of the subgroup analysis are available in Appendix 2. Concerning the results of subgroup analysis, 1) heterogeneity considerably reduced in subgroups compared to the main group, and 2) the absence of some subgroups in this category was due to less than two articles in that subgroup (Table 2).

**Table2.** Summary of subgroup analysis

| Outcome | Articles number | Sample size | HR | I<sup>2</sup> | Heterogeneity P value | CI (95%) |
| --- | --- | --- | --- | --- | --- | --- |
| **Overall survival** | **16** | **45099** | **2.24** | **56.1%** | **0.003** | **1.78-2.81** |
| FIGO 1988<sup>1</sup> | 4 | 732 | 1.97 | 42.7% | 0.155 | 1.23-3.16 |
| FIGO 2009 | 12 | 44530 | 2.36 | 61.6% | 0.003 | 1.80-3.10 |
| Type 1 histology | 7 | 43123 | 2.03 | 60.0% | 0.020 | 1.37-2.99 |
| Type 2 histology<sup>1</sup> | 3 | 472 | 2.73 | 20.7% | 0.283 | 1.56-4.77 |
| All histologies together<sup>1</sup> | 7 | 18518 | 2.15 | 50.5% | 0.059 | 1.60-2.89 |
| Low stages<sup>1</sup> | 7 | 43575 | 1.80 | 54.3% | 0.041 | 1.33-2.44 |
| High stages <sup>1</sup> | 3 | 338 | 1.92 | 47.6% | 0.149 | 1.22-3.01 |
| All stages together<sup>1</sup> | 8 | 2257 | 3.28 | 0.0% | 0.546 | 2.42-4.43 |
| **Disease free survival** | **18** | **6909** | **3.05** | **44.8%** | **0.021** | **2.35-3.97** |
| 1988 staging<sup>1</sup> | 3 | 422 | 1.59 | 0.0% | 0.553 | 1.10-2.30 |
| 2009 staging<sup>1</sup> | 15 | 6222 | 3.50 | 23.7% | 0.198 | 2.71-4.51 |
| Type 1 histology<sup>1</sup> | 6 | 3424 | 2.79 | 18.1% | 0.296 | 1.90-4.12 |
| Type 2 histology<sup>1</sup> | 4 | 514 | 5.04 | 0.0% | 0.418 | 2.82-9.01 |
| All histologies together | 9 | 2971 | 3.22 | 66.3% | 0.003 | 2.12-4.89 |
| Low stages | 8 | 3751 | 4.52 | 59.2% | 0.016 | 2.61-7.81 |
| High stages<sup>1</sup> | 4 | 530 | 1.87 | 0.0% | 0.658 | 1.36-2.57 |
| All stages together<sup>1</sup> | 9 | 3536 | 3.33 | 0.0% | 0.858 | 2.43-4.56 |
| **Cancer specific survival<sup>2</sup>** | **7** | **17920** | **3.46** | **81.7%** | **0.000** | **2.28-5.25** |
| 2009 staging | 6 | 17613 | 3.39 | 84.6% | 0.000 | 2.17-5.27 |
| Type 1 histology<sup>1</sup> | 4 | 27618 | 2.62 | 37.0% | 0.190 | 1.78-3.86 |
| All histologies together<sup>1</sup> | 4 | 15945 | 4.02 | 48.6% | 0.120 | 2.83-5.72 |
| Low stages | 4 | 42193 | 3.31 | 89.6% | 0.000 | 1.79-6.12 |
| All stages together<sup>1</sup> | 2 | 1045 | 4.38 | 70.9% | 0.064 | 1.68-11.40 |

## Discussion

Through this systematic review and meta-analysis, we demonstrated that positive peritoneal cytology can be used as an independent prognostic factor for endometrial cancer and it’s time to change back FIGO endometrial cancer staging. To guarantee our findings we also surveyed the prognostic value of PPC in endometrial carcinoma in 21 subgroups (based on tumor staging or histopathology), which all identified PPC as an independent prognostic factor for cancer of the endometrium.

Owing to rising technological and diagnostic technique advancements, the cancer staging system has been shown to be ever-changing, and endometrial cancers are no exception. FIGO staging for endometrial cancer was based on clinical evaluation until 1988, when it switched to pathological findings (7). In 2009, the system was also updated, with the removal of PC being one of the most significant changes (7). Many studies with extremely varied findings on the prognostic value of PPC in endometrial cancer have been published over a period of nearly 50 years (79). However, the prognostic value of PPC is still a matter of debate. Our findings in this systematic review and meta-analysis on 50 years of studies may be the end of this debate.

A study by Hirai et al. in 2001 evaluated the persistence of PPC on days 7 and 14 after peritoneal washing in patients with endometrial cancer with PPC. It was observed that only 5 out of 50 patients still had PPC on the 14th day. Although their findings questioned the prognostic value of PPC, they acknowledge that the prognostic role of PPC in endometrial cancer cannot be ignored (80).

In 2001, another study was carried out by Gürkan Arikan et al. in which, 24 patients with endometrial cancer and negative peritoneal cytology who underwent total abdominal hysterectomy were investigated and also some in vitro simulations were performed. The findings showed that cancerous cells in the peritoneal fluid were viable and were capable of implanting in the endometrial matrix (81).

The mechanisms of cancer cells migration and implantation seem to be taken into account. As it is evident from the terms used, the word “migration” indicates the movement of cells from one point to another which is different from the word “shedding”, and the positivity of PC can be intermittent (as three patients in Hirai’s study had negative PC on day 7 and PPC on day 14, while the author did not provide any convincing reason for this. However, it might be explained by cell migration or possibly cell alteration due to remaining in the peritoneal fluid). Furthermore, several studies have showed that tubal ligation during surgery can reduce the mortality rate in endometrial cancer patient.

Two other similar systematic reviews and meta-analyses have been conducted in years, and their findings are in line with ours (73)(82). However, in the present study, we examined the prognostic value of PC in EC patients with more enrolled articles, which was achieved by broadening the search strategy and minimizing inclusion and exclusion criteria with no time and language restrictions to increase the sensitivity of the search (by accepting the reduction of its specificity) and minimize selective reporting bias.

According to the results of the current study, 19 articles in total had concluded that PPC could not be considered as an independent prognostic factor. One of these studies which was carried out by Aiqin Wang et al. (2017) had the largest sample size among the reviewed articles (3415 patients, of whom 127 had PPC). They suggested that PPC, while being associated with other prognostic factors such as age and muscle invasion, could not be regarded as an independent prognostic factor, although it was identified that the risk of disease recurrence was significantly higher in patients with PPC compared to those with negative PC (χ2 = 7.970, P = 0.005).

There are a few limitations to our study that should be stated. Lack of considering the effects of treatment options on prognosis is one of them which can be reduced in future surveys with examining the patients in the same stages. Another drawback was that all studies included in our investigation were observational and retrospective type. Despite the mentioned limitations, the consistency of the subgroup analysis results enables us to consider the prognostic value of peritoneal cytology more confidently. Overall, it appears that more research with a larger sample size and higher quality is required to determine the prognostic value of peritoneal cytology. However, until more research is done, based on table 2 and the fact that the homogeneity of articles was acceptable in most subgroups, and all results in subgroups analysis were based on the positive effect of PC on the patient prognosis, as well as findings of previous systematic reviews on the subject, our findings suggest that PPC could be considered as an independent prognostic factor in patients with endometrial cancer. Therefore, it is recommended to be considered in the next FIGO Staging System for Endometrial Cancer to provide the patients with the necessary and sufficient treatments.

## Data Availability

All relevant data are within the manuscript and its Supporting Information files.
The data would be provided due to request from corresponding author.

## Declarations

## References

1. Goel JK, Kumar P, Malhotra N, Puri R. Cancer Endometrium: An Update. undefined. 2012;4(2):75–84.

2. Zhang S, Gong TT, Liu FH, Jiang YT, Sun H, Ma XX, et al. Global, Regional, and National Burden of Endometrial Cancer, 1990–2017: Results From the Global Burden of Disease Study, 2017. Front Oncol [Internet]. 2019 Dec 19 [cited 2022 Feb 1];9. Available from: /labs/pmc/articles/PMC6930915/

3. Bacalbasa N, Ionescu O. Clinic, pathologicand biologic prognostic factors in endometrial cancer. A literature review. Gineco.eu. 2015 Sep 1;11(3):134–7.

4. Makker V, Green AK, Wenham RM, Mutch D, Davidson B, Miller DS. New therapies for advanced, recurrent, and metastatic endometrial cancers. Gynecol Oncol Res Pract [Internet]. 2017 Dec [cited 2022 Feb 1];4(1). Available from: https://pubmed.ncbi.nlm.nih.gov/29214032/

5. Barak F, Kalichman L, Gdalevich M, Milgrom R, Laitman Y, Piura B, et al. The influence of early diagnosis of endometrioid endometrial cancer on disease stage and survival. Arch Gynecol Obstet [Internet]. 2013 Dec [cited 2022 Feb 1];288(6):1361–4. Available from: https://pubmed.ncbi.nlm.nih.gov/23695509/

6. Uharček P. Prognostic factors in endometrial carcinoma. J Obstet Gynaecol Res [Internet]. 2008 Oct [cited 2022 Feb 1];34(5):776–83. Available from: https://pubmed.ncbi.nlm.nih.gov/18958927/

7. Haltia UM, Bützow R, Leminen A, Loukovaara M. FIGO 1988 versus 2009 staging for endometrial carcinoma: a comparative study on prediction of survival and stage distribution according to histologic subtype. J Gynecol Oncol [Internet]. 2014 Jan [cited 2022 Feb 1];25(1):30–5. Available from: https://pubmed.ncbi.nlm.nih.gov/24459578/

8. Werner HMJ, Trovik J, Marcickiewicz J, Tingulstad S, Staff AC, Amant F, et al. Revision of FIGO surgical staging in 2009 for endometrial cancer validates to improve risk stratification. Gynecol Oncol [Internet]. 2012 Apr [cited 2022 Feb 1];125(1):103–8. Available from: https://pubmed.ncbi.nlm.nih.gov/22100838/

9. Gultekin M, Yildiz F, Ozyigit G, Beyaz H, Hayran M, Kose F, et al. Comparison of FIGO 1988 and 2009 staging systems for endometrial carcinoma. Med Oncol [Internet]. 2012 Dec [cited 2022 Feb 1];29(4):2955–62. Available from: https://pubmed.ncbi.nlm.nih.gov/22415398/

10. Lewin SN. Revised FIGO staging system for endometrial cancer. Clin Obstet Gynecol [Internet]. 2011 Jun [cited 2022 Feb 1];54(2):215–8. Available from: https://journals.lww.com/clinicalobgyn/Fulltext/2011/06000/Revised_FIGO_Staging_System_for_Endometrial_Cancer.4.aspx

11. Seagle BLL, Alexander AL, Lantsman T, Shahabi S. Prognosis and treatment of positive peritoneal cytology in early endometrial cancer: matched cohort analyses from the National Cancer Database. Am J Obstet Gynecol [Internet]. 2018 Mar 1 [cited 2022 Feb 1];218(3):329.e1-329.e15. Available from: https://pubmed.ncbi.nlm.nih.gov/29223598/

12. Garg G, Gao F, Wright JD, Hagemann AR, Mutch DG, Powell MA. Positive peritoneal cytology is an independent risk-factor in early stage endometrial cancer. Gynecol Oncol [Internet]. 2013 Jan [cited 2022 Feb 1];128(1):77–82. Available from: https://pubmed.ncbi.nlm.nih.gov/23032094/

13. Milgrom SA, Kollmeier MA, Abu-Rustum NR, Makker V, Gardner GJ, Barakat RR, et al. Positive peritoneal cytology is highly predictive of prognosis and relapse patterns in stage III (FIGO 2009) endometrial cancer. Gynecol Oncol [Internet]. 2013 Jul [cited 2022 Feb 1];130(1):49–53. Available from: https://pubmed.ncbi.nlm.nih.gov/23603151/

14. Shiozaki T, Tabata T, Yamada T, Yamamoto Y, Yamawaki T, Ikeda T. Does positive peritoneal cytology not affect the prognosis for stage I uterine endometrial cancer?: the remaining controversy and review of the literature. Int J Gynecol Cancer [Internet]. 2014 [cited 2022 Feb 1];24(3):549–55. Available from: https://pubmed.ncbi.nlm.nih.gov/24476896/

15. Moher D, Liberati A, Tetzlaff J, Altman DG, Altman D, Antes G, et al. Preferred reporting items for systematic reviews and meta-analyses: the PRISMA statement. PLoS Med [Internet]. 2009 Jul [cited 2022 Feb 6];6(7). Available from: https://pubmed.ncbi.nlm.nih.gov/19621072/

16. Binesh F, Moravej MT, Heydari S, Aghabagheri M, Dehghan H. Evaluation of the Prognostic Value of Peritoneal Cytology in Patients with Endometrial Cancer: A Protocol for a Systematic Review and Meta-Analysis. Crit Comments Biomed [Internet]. 2021 Mar 1 [cited 2022 Feb 6];2(1):100–3. Available from: http://ccbjournal.ssu.ac.ir/article_106.html

17. Hayden JA, Côté P, Bombardier C. Evaluation of the quality of prognosis studies in systematic reviews. Ann Intern Med [Internet]. 2006 Mar 21 [cited 2022 Feb 8];144(6):427–37. Available from: https://pubmed.ncbi.nlm.nih.gov/16549855/

18. Hayden JA, van der Windt DA, Cartwright JL, Côté P, Bombardier C. Assessing bias in studies of prognostic factors. Ann Intern Med [Internet]. 2013 Feb 19 [cited 2022 Feb 8];158(4):280–6. Available from: https://pubmed.ncbi.nlm.nih.gov/23420236/

19. Cetinkaya K, Atalay F. Peritoneal cytology in endometrial cancer. Tumori [Internet]. 2015 Nov 1 [cited 2022 Mar 9];101(6):697–700. Available from: https://pubmed.ncbi.nlm.nih.gov/26350193/

20. Sutton GP, Geisler HE, Stehman FB, Young PCM, Kimes TM, Ehrlich CE. Features associated with survival and disease-free survival in early endometrial cancer. Am J Obstet Gynecol [Internet]. 1989 [cited 2022 Mar 9];160(6):1385–93. Available from: https://pubmed.ncbi.nlm.nih.gov/2735364/

21. Comparative Survival Outcomes of Uterine Papillary Serous Carcinoma, Clear Cell Carcinoma, Grade 3 Endometrioid Adenocarcinoma, and Carcinosarcoma of Endometrial Cancer in Rajavithi Hospital - PubMed [Internet]. [cited 2022 Mar 9]. Available from: https://pubmed.ncbi.nlm.nih.gov/27266220/

22. Tang X, Tanemura K, Ye W, Ohmi K, Tsunematsu R, Yamada T, et al. Clinicopathological factors predicting retroperitoneal lymph node metastasis and survival in endometrial cancer. Jpn J Clin Oncol [Internet]. 1998 [cited 2022 Mar 9];28(11):673–8. Available from: https://pubmed.ncbi.nlm.nih.gov/9861234/

23. Grigsby PW, Perez CA, Kuten A, Simpson JR, Garcia DM, Camel HM, et al. Clinical stage I endometrial cancer: prognostic factors for local control and distant metastasis and implications of the new FIGO surgical staging system. Int J Radiat Oncol Biol Phys [Internet]. 1992 [cited 2022 Mar 9];22(5):905–11. Available from: https://pubmed.ncbi.nlm.nih.gov/1555983/

24. Hanley KZ, Fadare O, Fisher KE, Atkins KA, Mosunjac MB. Clinical Significance of Positive Pelvic Washings in Uterine Papillary Serous Carcinoma Confined to an Endometrial Polyp. Int J Gynecol Pathol [Internet]. 2016 [cited 2022 Mar 9];35(3):249–55. Available from: https://pubmed.ncbi.nlm.nih.gov/26535985/

25. Sasada S, Yunokawa M, Takehara Y, Ishikawa M, Ikeda S, Kato T, et al. Baseline risk of recurrence in stage I–II endometrial carcinoma. J Gynecol Oncol [Internet]. 2018 Jan 1 [cited 2022 Mar 9];29(1). Available from: /labs/pmc/articles/PMC5709535/

26. Mariani A, Webb MJ, Keeney GL, Aletti G, Podratz KC. Assessment of prognostic factors in stage IIIA endometrial cancer. Gynecol Oncol [Internet]. 2002 [cited 2022 Mar 9];86(1):38–44. Available from: https://pubmed.ncbi.nlm.nih.gov/12079298/

27. Kulhan M, Kulhan G, Nayki U, Nayki C, Ulug P, Sipahi M, et al. Assessment of clinicopathological features, evaluation of treatment, and prognosis of clear cell and serous papillary endometrial carcinoma. Ginekol Pol [Internet]. 2016 [cited 2022 Mar 9];87(8):570–4. Available from: https://pubmed.ncbi.nlm.nih.gov/27629131/

28. McMeekin DS, Lashbrook D, Gold M, Johnson G, Walker JL, Mannel R. Analysis of FIGO Stage IIIc endometrial cancer patients. Gynecol Oncol [Internet]. 2001 [cited 2022 Mar 9];81(2):273–8. Available from: https://pubmed.ncbi.nlm.nih.gov/11330962/

29. Konski A, Poulter C, Keys H, Rubin P, Beecham J, Doane K. Absence of prognostic significance, peritoneal dissemination and treatment advantage in endometrial cancer patients with positive peritoneal cytology. Int J Radiat Oncol Biol Phys [Internet]. 1988 Jan 11 [cited 2022 Mar 9];14(1):49–55. Available from: https://eurekamag.com/research/004/648/004648457.php

30. Multivariate analysis of prognostic factors in patients with endometrial cancer - PubMed [Internet]. [cited 2022 Mar 9]. Available from: https://pubmed.ncbi.nlm.nih.gov/8737300/

31. Yazigi R, Piver M, gynecology LB-O and, 1983 undefined. Malignant peritoneal cytology as prognostic indicator in stage I endometrial cancer. http://europepmc.org [Internet]. [cited 2022 Mar 9]; Available from: https://europepmc.org/article/med/6877692

32. Gal D, Recio FO, Zamurovic D, Tancer ML. Lymphvascular space involvement—A prognostic indicator in endometrial adenocarcinoma. Gynecol Oncol. 1991 Aug 1;42(2):142–5.

33. Saga Y, Imai M, Jobo T, Kuramoto H, Takahashi K, Konno R, et al. Is peritoneal cytology a prognostic factor of endometrial cancer confined to the uterus? Gynecol Oncol [Internet]. 2006 Oct [cited 2022 Mar 9];103(1):277–80. Available from: https://pubmed.ncbi.nlm.nih.gov/16678244/

34. Preyer O, Obermair A, Formann E, Schmid W, Perrin LC, Ward BG, et al. The impact of positive peritoneal washings and serosal and adnexal involvement on survival in patients with stage IIIA uterine cancer. Gynecol Oncol [Internet]. 2002 [cited 2022 Mar 9];86(3):269–73. Available from: https://pubmed.ncbi.nlm.nih.gov/12217747/

35. Tanaka K, Kobayashi Y, Sugiyama J, Yamazaki T, Dozono K, Watanabe M, et al. Histologic grade and peritoneal cytology as prognostic factors in type 1 endometrial cancer. Int J Clin Oncol [Internet]. 2017 Jun 1 [cited 2022 Mar 9];22(3):533–40. Available from: https://pubmed.ncbi.nlm.nih.gov/28083737/

36. Hoekstra A V., Kim RJ, Small W, Rademaker AW, Helenowski IB, Singh DK, et al. FIGO stage IIIC endometrial carcinoma: prognostic factors and outcomes. Gynecol Oncol [Internet]. 2009 Aug [cited 2022 Mar 9];114(2):273–8. Available from: https://pubmed.ncbi.nlm.nih.gov/19428094/

37. Zuna RE, Behrens A. Peritoneal washing cytology in gynecologic cancers: long-term follow-up of 355 patients. J Natl Cancer Inst [Internet]. 1996 Jul 17 [cited 2022 Mar 9];88(14):980–7. Available from: https://pubmed.ncbi.nlm.nih.gov/8667429/

38. Obermair A, Geramou M, Tripcony L, Nicklin JL, Perrin L, Crandon AJ. Peritoneal cytology: impact on disease-free survival in clinical stage I endometrioid adenocarcinoma of the uterus. Cancer Lett [Internet]. 2001 Mar 10 [cited 2022 Mar 9];164(1):105–10. Available from: https://pubmed.ncbi.nlm.nih.gov/11166922/

39. Han KH, Park NH, Kim HS, Chung HH, Kim JW, Song YS. Peritoneal cytology: a risk factor of recurrence for non-endometrioid endometrial cancer. Gynecol Oncol [Internet]. 2014 [cited 2022 Mar 9];134(2):293–6. Available from: https://pubmed.ncbi.nlm.nih.gov/24854062/

40. Imachi M, Tsukamoto N, Matsuyama T, Nakano H. Peritoneal cytology in patients with endometrial carcinoma. Gynecol Oncol. 1988 May 1;30(1):76–86.

41. Ito K, Inoue Y, Obata K, of KN-TTJ, 1992 undefined. Peritoneal cytology in endometrial carcinoma. jstage.jst.go.jp [Internet]. 1992 [cited 2022 Mar 9];166(3):387–94. Available from: https://www.jstage.jst.go.jp/article/tjem1920/166/3/166_3_387/_article/-char/ja/

42. Atalay F, Çetinkaya K. Uterin Papiller Seröz Kanserde Prognostik Faktörler: Ankara Onkoloji Hastanesi Deneyimi. J Clin Obstet Gynecol [Internet]. 2015 Jul 14 [cited 2022 Mar 9];25(3):168–72. Available from: https://www.jcog.com.tr/article/prognostic-factors-in-uterine-papillary-serous-cancer-ankara-oncology-hospital-experience-71503.html

43. Tate K, Yoshida H, Ishikawa M, Uehara T, Ikeda SI, Hiraoka N, et al. Prognostic factors for patients with early-stage uterine serous carcinoma without adjuvant therapy. J Gynecol Oncol [Internet]. 2018 May 1 [cited 2022 Mar 9];29(3). Available from: https://pubmed.ncbi.nlm.nih.gov/29533019/

44. Milgrom SA, Kollmeier MA, Abu-Rustum NR, Makker V, Gardner GJ, Barakat RR, et al. Positive peritoneal cytology is highly predictive of prognosis and relapse patterns in stage III (FIGO 2009) endometrial cancer. Gynecol Oncol [Internet]. 2013 Jul [cited 2022 Mar 9];130(1):49–53. Available from: https://pubmed.ncbi.nlm.nih.gov/23603151/

45. Garg G, Gao F, Wright JD, Hagemann AR, Mutch DG, Powell MA. Positive peritoneal cytology is an independent risk-factor in early stage endometrial cancer. Gynecol Oncol [Internet]. 2013 Jan [cited 2022 Mar 9];128(1):77–82. Available from: https://pubmed.ncbi.nlm.nih.gov/23032094/

46. Kadar N, Homesley HD, Malfetano JH. Positive peritoneal cytology is an adverse factor in endometrial carcinoma only if there is other evidence of extrauterine disease. Gynecol Oncol [Internet]. 1992 [cited 2022 Mar 9];46(2):145–9. Available from: https://pubmed.ncbi.nlm.nih.gov/1500014/

47. Takeshima N, Nishida H, Tabata T, Hirai Y, Hasumi K. Positive peritoneal cytology in endometrial cancer: enhancement of other prognostic indicators. Gynecol Oncol [Internet]. 2001 [cited 2022 Mar 9];82(3):470–3. Available from: https://pubmed.ncbi.nlm.nih.gov/11520142/

48. Akhavan S, Kazemi Z, Alibakhshi A, Modaresgilani M, Mousavi A, Ahmadzadeh A, et al. Positive Peritoneal Cytology as a Predictor of Prognosis in Early Stage of Endometrioid Adenocarcinoma. Iran J Cancer Prev 2017 102 [Internet]. 2017 Feb 28 [cited 2022 Mar 9];10(2). Available from: https://brief.land/ijcm/articles/5285.html

49. Scott SA, van der Zanden C, Cai E, McGahan CE, Kwon JS. Prognostic significance of peritoneal cytology in low-intermediate risk endometrial cancer. Gynecol Oncol [Internet]. 2017 May 1 [cited 2022 Mar 9];145(2):262–8. Available from: https://pubmed.ncbi.nlm.nih.gov/28359690/

50. Tebeu PM, Popowski Y, Verkooijen HM, Bouchardy C, Ludicke F, Usel M, et al. Positive peritoneal cytology in early-stage endometrial cancer does not influence prognosis. Br J Cancer 2004 914 [Internet]. 2004 Jul 13 [cited 2022 Mar 9];91(4):720–4. Available from: https://www.nature.com/articles/6602035

51. Risk factors of cytologic positive rate of ascites in early endometrial cancer patients and its influences on prognosis | Abstract [Internet]. [cited 2022 Mar 9]. Available from: https://www.alliedacademies.org/abstract/risk-factors-of-cytologic-positive-rate-of-ascites-in-early-endometrial-cancer-patients-and-its-influences-on-prognosis-8837.html

52. Türkmen O, Karalok A, Başaran D, Kimyon G, Kul G, Tulunay G, et al. Revaluating the survival effects of International Federation of Gynecology and Obstetrics 1988 stage IIIA criteria for endometrial cancer. J Turkish-German Gynecol Assoc [Internet]. 2017 Sep 1 [cited 2022 Mar 9];18(3):110–5. Available from: https://app.trdizin.gov.tr/makale/TWpnM01qYzNOdz09/revaluating-the-survival-effects-of-international-federation-of-gynecology-and-obstetrics-1988-stage-iiia-criteria-for-endometrial-cancer

53. Milgrom SA, Kollmeier MA, Abu-Rustum NR, O’Cearbhaill RE, Barakat RR, Alektiar KM. Quantifying the risk of recurrence and death in stage III (FIGO 2009) endometrial cancer. Gynecol Oncol [Internet]. 2014 [cited 2022 Mar 9];134(2):297–301. Available from: https://pubmed.ncbi.nlm.nih.gov/24875123/

54. Binesh F, Akhavan A, Behniafard N, Zabihi S, Hosseinizadeh E. Prognostic value of peritoneal washing cytology in gynecologic malignancies: a controversial issue. Asian Pac J Cancer Prev [Internet]. 2014 [cited 2022 Mar 9];15(21):9405–10. Available from: https://pubmed.ncbi.nlm.nih.gov/25422232/

55. Ide P. Prognostic value of peritoneal fluid cytology in patients with endometrial cancer stage I. Eur J Obstet Gynecol Reprod Biol [Internet]. 1984 [cited 2022 Mar 9];18(5–6):343–9. Available from: https://pubmed.ncbi.nlm.nih.gov/6526121/

56. Prognostic value of cytologic examination of peritoneal washings in patients with endometrial carcinoma - PubMed [Internet]. [cited 2022 Mar 9]. Available from: https://pubmed.ncbi.nlm.nih.gov/6171978/

57. Grimshaw RN, Carl Tupper W, Fraser RC, Tompkins MG, Jeffrey JF. Prognostic value of peritoneal cytology in endometrial carcinoma. Gynecol Oncol [Internet]. 1990 [cited 2022 Mar 9];36(1):97–100. Available from: https://pubmed.ncbi.nlm.nih.gov/2295459/

58. Kasamatsu T, Onda T, Katsumata N, Sawada H, Yamada T, Tsunematsu R, et al. Prognostic significance of positive peritoneal cytology in endometrial carcinoma confined to the uterus. Br J Cancer 2003 882 [Internet]. 2003 Jan 28 [cited 2022 Mar 9];88(2):245–50. Available from: https://www.nature.com/articles/6600698

59. Mazurka JL, Krepart G V., Lotocki RJ. Prognostic significance of positive peritoneal cytology in endometrial carcinoma. Am J Obstet Gynecol [Internet]. 1988 [cited 2022 Mar 9];158(2):303–6. Available from: https://pubmed.ncbi.nlm.nih.gov/3341409/

60. Havrilesky LJ, Secord AA, O’Malley DM, Broadwater G, Bae-Jump V, Cohn DE, et al. Multicenter analysis of recurrence and survival in stage IIIA endometrial cancer. Gynecol Oncol [Internet]. 2009 Aug [cited 2022 Mar 9];114(2):279–83. Available from: https://pubmed.ncbi.nlm.nih.gov/19446319/

61. Lo SST, Cheng DKL, Ng TY, Wong LC, Ngan HYS. The prognostic significance of positive peritoneal cytology in endometrial cancer. Int J Gynecol Cancer. 1996;6(6):473–6.

62. Havrilesky LJ, Cragun JM, Calingaert B, Alvarez Secord A, Valea FA, Clarke-Pearson DL, et al. The prognostic significance of positive peritoneal cytology and adnexal/serosal metastasis in stage IIIA endometrial cancer. Gynecol Oncol [Internet]. 2007 Feb [cited 2022 Mar 9];104(2):401–5. Available from: https://pubmed.ncbi.nlm.nih.gov/17014898/

63. Creasman WT, Disaia PJ, Blessing J, Wilkinson RH, Johnston W, Weed JC. Prognostic significance of peritoneal cytology in patients with endometrial cancer and preliminary data concerning therapy with intraperitoneal radiopharmaceuticals. Am J Obstet Gynecol. 1981 Dec 15;141(8):921–9.

64. Matsuo K, Yabuno A, Hom MS, Shida M, Kakuda M, Adachi S, et al. Significance of abnormal peritoneal cytology on survival of women with stage I-II endometrioid endometrial cancer. Gynecol Oncol [Internet]. 2018 May 1 [cited 2022 Mar 9];149(2):301–9. Available from: https://pubmed.ncbi.nlm.nih.gov/29605499/

65. Kyrgiou M, Chatterjee J, Lyus R, Amin T, Ghaem-Maghami S. The role of cytology and other prognostic factors in endometrial cancer. https://doi.org/103109/014436152013813916 [Internet]. 2013 Oct [cited 2022 Mar 9];33(7):729–34. Available from: https://www.tandfonline.com/doi/abs/10.3109/01443615.2013.813916

66. Kashimura M, Sugihara K, Toki N, Matsuura Y, Kawagoe T, Kamura T, et al. The significance of peritoneal cytology in uterine cervix and endometrial cancer. Gynecol Oncol [Internet]. 1997 [cited 2022 Mar 9];67(3):285–90. Available from: https://pubmed.ncbi.nlm.nih.gov/9441776/

67. The prognostic significance of peritoneal cytology for stage I endometrial cancer - PubMed [Internet]. [cited 2022 Mar 9]. Available from: https://pubmed.ncbi.nlm.nih.gov/2812655/

68. Vizza E, Mancini E, Laquintana V, Loria R, Carosi M, Baiocco E, et al. The prognostic significance of positive peritoneal cytology in endometrial cancer and its correlations with L1-CAM biomarker. Surg Oncol [Internet]. 2019 Mar 1 [cited 2022 Mar 9];28:151–7. Available from: https://pubmed.ncbi.nlm.nih.gov/30851892/

69. Yildirim N, Bilgi A, Gokulu SG, Akman L, Zekioglu O, Serin G, et al. Lymphovascular space invasion and positive peritoneal cytology are independent prognostic factors for lymph node metastasis and recurrence in endometrial cancer. Eur J Gynaecol Oncol [Internet]. 2018 Dec 10 [cited 2022 Mar 9];39(6):977–83. Available from: https://www.imrpress.com/journal/EJGO/39/6/10.12892/ejgo4539.2018

70. Dong Y, Wang Z, Wang J. Positive peritoneal cytology is an independent risk factor in endometrial cancer. J Obstet Gynaecol Res [Internet]. 2020 Sep 1 [cited 2022 Mar 9];46(9):1842–50. Available from: https://pubmed.ncbi.nlm.nih.gov/32643298/

71. Fadare O, Mariappan MR, Hileeto D, Wang S, McAlpine JN, Rimm DL. Upstaging based solely on positive peritoneal washing does not affect outcome in endometrial cancer. Mod Pathol [Internet]. 2005 May [cited 2022 Mar 9];18(5):673–80. Available from: https://pubmed.ncbi.nlm.nih.gov/15578078/

72. Takahashi K, Yunokawa M, Sasada S, Takehara Y, Miyasaka N, Kato T, et al. A novel prediction score for predicting the baseline risk of recurrence of stage I–II endometrial carcinoma. J Gynecol Oncol [Internet]. 2019 Jan 1 [cited 2022 Mar 9];30(1). Available from: /labs/pmc/articles/PMC6304400/

73. Matsuo K, Matsuzaki S, Nusbaum DJ, Machida H, Nagase Y, Grubbs BH, et al. Malignant peritoneal cytology and decreased survival of women with stage I endometrioid endometrial cancer. Eur J Cancer [Internet]. 2020 Jul 1 [cited 2022 Mar 9];133:33–46. Available from: https://pubmed.ncbi.nlm.nih.gov/32434109/

74. Cheng Y, Dong Y, Tian W, Zhang H, Li X, Wang Z, et al. Nomogram for Predicting Recurrence-Free Survival in Chinese Women with Endometrial Cancer after Initial Therapy: External Validation. J Oncol. 2020;2020.

75. Wang L, Li L, Wu M, Lang J. The prognostic role of peritoneal cytology in stage IA endometrial endometrioid carcinomas. Curr Probl Cancer [Internet]. 2020 Apr 1 [cited 2022 Mar 9];44(2). Available from: https://pubmed.ncbi.nlm.nih.gov/31753315/

76. Bezircioglu I, Yetimalar MH, Kilic D. Factors affecting the prognosis and overall survival in patients with uterine papillary serous carcinoma. J Clin Diagnostic Res. 2018 May 1;12(5):QC01–5.

77. Zhong X, Wang J, Kaku T, Wang Z, Li X, Wei L. Prognostic Factors of Uterine Serous Carcinoma-A Multicenter Study. Int J Gynecol Cancer [Internet]. 2018 Jul 1 [cited 2022 Mar 9];28(6):1138–44. Available from: https://pubmed.ncbi.nlm.nih.gov/29621127/

78. Santala M, Talvensaari-Mattila A, research AK-A, 2003 undefined. Peritoneal cytology and preoperative serum CA 125 level are important prognostic indicators of overall survival in advanced endometrial cancer. http://europepmc.org [Internet]. [cited 2022 Apr 17]; Available from: https://europepmc.org/article/med/12926169

79. Binesh F, Akhavan A, Behniafard N, Zabihi S, Hosseinizadeh E. Prognostic value of peritoneal washing cytology in gynecologic malignancies: a controversial issue. Asian Pac J Cancer Prev [Internet]. 2014 [cited 2022 Feb 20];15(21):9405–10. Available from: https://pubmed.ncbi.nlm.nih.gov/25422232/

80. Y H, N T, T K, K H. Malignant potential of positive peritoneal cytology in endometrial cancer. Obstet Gynecol [Internet]. 2001 May [cited 2022 Feb 20];97(5 Pt 1):725–8. Available from: https://pubmed.ncbi.nlm.nih.gov/11339924/

81. Arikan G, Reich O, Weiss U, Hahn T, Reinisch S, Tamussino K, et al. Are endometrial carcinoma cells disseminated at hysteroscopy functionally viable? Gynecol Oncol [Internet]. 2001 [cited 2022 Mar 9];83(2):221–6. Available from: https://pubmed.ncbi.nlm.nih.gov/11606075/

82. Lee B, Suh DH, Kim K, No JH, Kim YB. Influence of positive peritoneal cytology on prognostic factors and survival in early-stage endometrial cancer: a systematic review and meta-analysis. Jpn J Clin Oncol [Internet]. 2016 Aug 1 [cited 2022 Feb 20];46(8):711–7. Available from: https://pubmed.ncbi.nlm.nih.gov/27207885/

